# COVID-19 Outbreak in Post-Soviet States: Modeling the Best and Worst Possible Scenarios

**DOI:** 10.1101/2020.04.19.20071704

**Authors:** Alpamys Issanov, Yerlan Amanbek, Anara Abbay, Shalkar Adambekov, Mohamad Aljofan, Ardak Kashkynbayev, Abduzhappar Gaipov

**Affiliations:** Department of Clinical Sciences, Nazarbayev University School of Medicine, Nur-Sultan, Kazakhstan; Department of Mathematics, Nazarbayev University School of Sciences and Humanities, Nur-Sultan, Kazakhstan; Department of Epidemiology, University of Pittsburgh Graduate School of Public Health, Pittsburgh, PA, USA; Department of Biomedical Sciences, Nazarbayev University School of Medicine, Nur-Sultan, Kazakhstan

**Keywords:** COVID-19, Modelling, Central Asia, Post-Soviet states, pandemic, scenarios

## Abstract

**Background:** COVID-19 pandemic has presented extreme challenges to developing countries across the world. Post-Soviet states are facing unique challenges due to their developing healthcare systems and unstable economy. The aim of this paper was to provide estimates for current development COVID-19 pandemic in the Post-Soviet states and forecast potential best and worst scenarios for spread of this deadly infection.

**Methods:** The data on confirmed cases and deaths were extracted from official governmental sources for a period from beginning of outbreak dates for each country until April 18, 2020. A modified SEIR (Susceptible-Exposed-Infected-Recovered) modelling was used to plot the parameters of epidemic in 10 post-Soviet states and forecast the number of cases over a period of 10, 30 and 60 days. We also estimated the numbers of cases based on the optimal measures (best scenario) and suboptimal measures (worst scenarios) of potential spread of COVID-19 in these countries.

**Results:** It was estimated that Armenia and Azerbaijan have reached their peaks, Kazakhstan, Kyrgyzstan, Moldova and Uzbekistan are expected to reach their peaks in the coming week (April 29 – May 7, 2020), with comparatively low cases of COVID-19 and loss of lives in the best-case scenario. In contrast, Belarus, Russia, and Ukraine would likely see the outbreaks with the largest number of COVID-19 cases amongst the studied Post-Soviet States in the worst scenario during the next 30 and 60 days. Geographical remoteness and small number of international travelers from the countries heavily affected by the pandemic could also have contributed to delay in the spread of COVID-19.

**Conclusion:** Governmental response was shown to be as an important determining factor responsible for the development of COVID-19 epidemic in Post-Soviet states. The current protection rates should be maintained to reduce active cases during upcoming 30 and 60 days. The estimated possible scenarios based on the proposed model can potentially be used by healthcare professionals from each studied Post-Soviet States as well as others to improve plans to contain the current and future epidemic.

## INTRODUCTION

The recent pandemic of acute respiratory tract infection caused by a novel strain of coronavirus originated in Wuhan city, China, in December 2019 (1, 2). The International Committee of Taxonomy of Viruses officially named the novel coronavirus as “Severe acute respiratory syndrome coronavirus 2 (SARS-CoV-2)”; and the World Health Organization (WHO) labelled the clinical syndromes caused by SARS-CoV-2 as Coronavirus disease (COVID-19) (3-5). The virus spreads rapidly across the globe affecting 210 countries and territories with over 2.2M confirmed cases and total 155K confirmed deaths as of April 17th, 2020 (4). Europe (Italy, Spain, France, Germany), USA, UK, China and Iran have reported the highest incidence and mortality from COVID-19 leading to catastrophic health, economic and social repercussions (4). COVID-19 is expected to have even more severe impact on developing countries once the pandemic reaches its peak in regions with more vulnerable economies and health systems.

These include the healthcare systems in the Post-Soviet states, which are transitioning from the Soviet healthcare system, characterized by a centralized system of governance of hospitals, public health organizations, and health departments, to modernized healthcare systems adaptable to market economy (6). These include three developing countries in the Central Asia region and seven countries from Eastern European region, which for the best part of the 20th century, were part of the Soviet Union until their independence in the early 1990s. While these counties currently have different economic and political systems, most of these countries have similar healthcare systems and public health challenges inherited from Soviet Union (7). However, the transitioning seems to be slower than expected and that these systems remain somewhat unchanged from the Soviet era, highlighting the need for rapid improvement of resources and patient access.

Following the confirmation of the first of COVID-19 cases in a part of Europe, most of the Post-Soviet states took various pathways in introducing and tightening preventive measures of COVID-19 outbreaks, such as limiting or suspending all public transportation, cancelling of public events, restricting people from leaving their residence, and complete lockdown of cities (8-13). An outbreak predicting system is an extremely useful and vital tool of preparedness of such events. The majority of these systems utilizes available data and applies mathematical and/or statistical modeling capable of predicting a future outbreak (14, 15). There were several attempts in different countries to establish a modelling method for the prediction of COVID-19 spread including a group in China which estimated the serial interval for COVID-19 (16), to predict the dynamics of COVID-19 spread (17). Applying a mathematical model on the available data, a group showed that lockdown measures were effective in reducing COVID-19 transmission rate and that imported cases have a different dynamic of transmission. Similarly, using an age-structured compartmental model of COVID-19 transmission, a Canadian group showed that significant strengthening of quarantine measures could prevent extreme overloading of intensive care unit (ICU) resources (18). However, the situation in the Post-Soviet countries remains uninvestigated.

To date, each the Post-Soviet countries initiated their own preventive actions against the spread of COVID-19, which led to high healthcare expenditures and economic slow-down (13). The high economic, social and health impacts of the pandemic make it hard to determine how long each of these developing countries would be able to maintain the strict measures. Therefore, there is a need for a reliable prediction model for these countries, to determine the status of preparedness. A system capable of determining whether governmental preparedness policies are maintained, and the spread is stopped/slowed as a best-case scenario; or policies not maintained and thus disease spread continuously unabated, worst case scenario. Such models are important to provide the projected estimates that can be used by governments and public health practitioners who are responsible for responding to COVID-19 epidemic.

Therefore, the aim of this study was to provide a reliable mathematical model capable of determining the best- and worst-case scenarios for the development of COVID-19 epidemic in the Post-Soviet States, considering the effects of optimal and suboptimal measures, respectively. The model is based on a modified version of the Susceptible-Exposed-Infectious-Recovered (SEIR) deterministic mathematical model. We will also discuss how the measures taken by each country are affecting the current situation in Post-Soviet States, as well as providing a predictive scenario of the best- and worst-cases for this highly vulnerable region in comparison with other Post-Soviet States.

## METHODS

### Data collection

Data on country-specific population size, population density and percentage of population 65 years or over were extracted from the United Nations (UN) official website (19, 20). Health expenditure and country ranking data were collected from the World Bank databases (21). The number of air passengers carried in both domestic and international flights for each country were obtained from the International Civil Aviation Organization, except Armenia whose data was collected from the International Air Transport Association (22, 23).

The total daily data of newly confirmed COVID-19 infected, recovered cases and number of COVID-19 deaths in selected Post-Soviet States in the region were obtained from the official websites of the Ministries of Health in each of the studied countries including Armenia, Azerbaijan, Belarus, Georgia, Kazakhstan, Kyrgyzstan, Moldova, Russia, Ukraine, and Uzbekistan (13). Given the absence of COVID-19 infected cases, Tajikistan and Turkmenistan were excluded from the study. Also, Baltic states (Estonia, Latvia and Lithuania) were excluded due to the difference in their health care systems, policy implementation and economic situation to that of the Post-Soviet States. Of note, the included COVID-19 cases were laboratory confirmed using WHO guidelines (24) while recovered cases were defined as “those that were previously tested positive to COVID-19 (laboratory confirmed) and later had negative test results”. While some of the studied countries introduced serological testing to investigate retrospective outbreaks by identifying asymptomatic immunized people, they were excluded in the current mathematical model of forecasting.

### Mathematical modeling method

The two most common deterministic models used in the literature are the logistic and modified Susceptible-Infectious-Recovered (SIR) models (15, 17). Although the logistic model requires less data, it underestimates peak timing and the number of cases (25). Instead, a modified SEIR model “Susceptible-Protected-Exposed-Infectious-Quarantined-Recovered-Dead” (SPEIQRD) was used (26). This SPEIQRD framework incorporates additional public health interventions such as, self-isolation of exposed, quarantine of infectious and isolation of susceptible. Parameters describing the natural history and clinical path of COVID-19 were derived from published literature. An overview of the SPEIQRD framework, its compartments and movements between them can be found in **Figure 1**. More information about each compartment can be found in Supplementary data (**Appendix 1, Model 1**). The forecasting model was performed to cover a period of 10, 30 and 60 days from April 18, 2020, and it was assumed that recovered people remain immune from reinfection for the duration of the pandemic. The model assumes that individuals remained infectious until they recovered, quarantined, or died and that all confirmed cases would have been quarantined. In the worst case scenario, it was assumed that the rate of movement from susceptible to protected (protection rate, alpha) will decrease by 50%, whereas in the best case scenario, the protection rate increases by 20% as suggested by the literature (18). The model was performed in Matlab 19a version (The MathWorks, Inc., Natick, MA 01760, USA).

**Figure 1.**
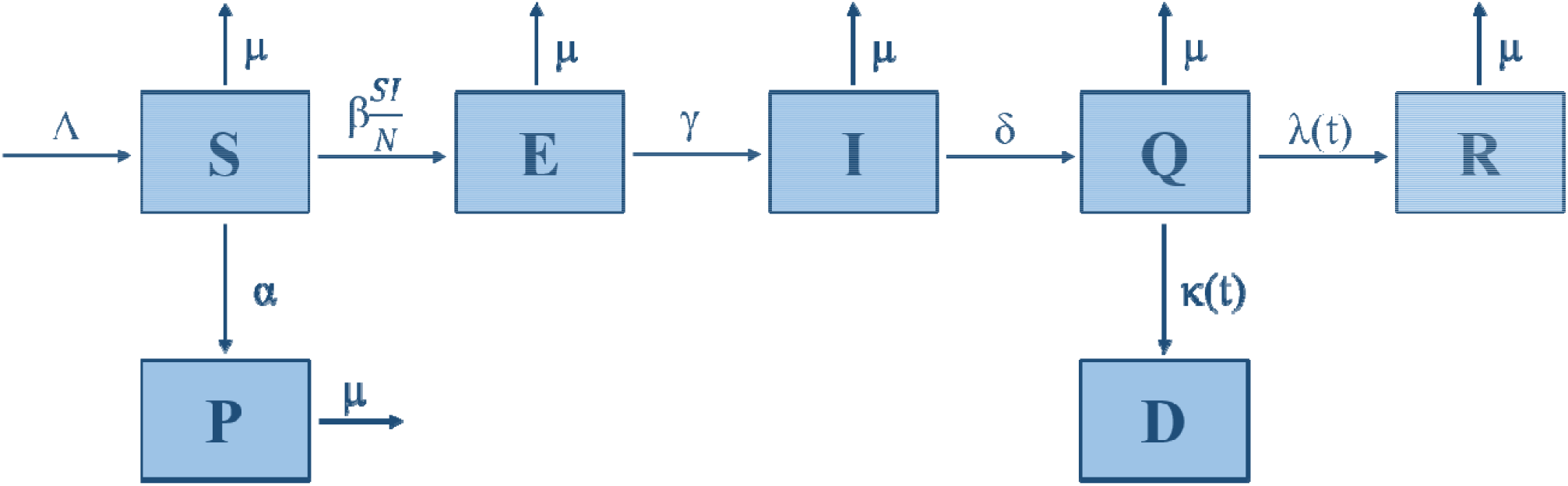
Algorithm of SPEIQRD model.

### Numerical Simulations

For the numerical simulations, we ignored the birth and natural death rates since they have insignificant effect compared to other parameters such as the high transmission rate of COVID-19. That is, we set A= 0 and µ = 0 and we modify the model as follows in (**Model 2**), which was proposed by Peng, et al (2020) [1]. We have estimated parameters by fitting to available COVID-19 data from the official websites of Ministry of Health in each country. The function for the fitting is Matlab’s function lsqcurvefit (27) and the code was derived from E. Cheynet et al. (28). Modified SPIERQD **Model 2** presented in following equations:

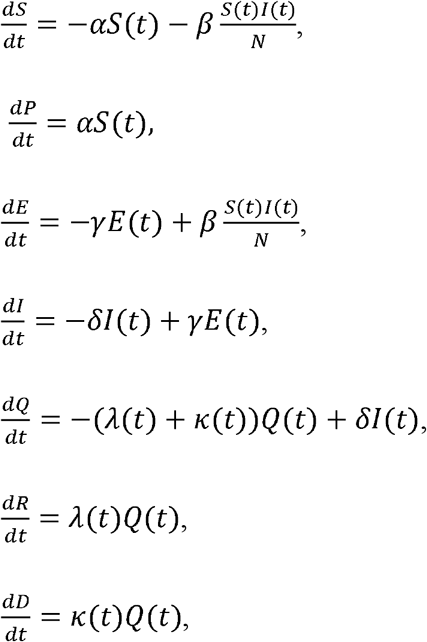

One can easily see that for the modified model the basic reproduction number simplifies to

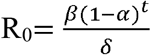

where t represents number of days.

Based on above calculations, the reproduction number (R_0_) for each country was generated and presented in **Supplementary Table 1** as well as dynamical changes of R_0_ over the time depicted in **Supplementary Figure 1**.

## RESULTS

### Country specific information and healthcare expenditure data

General country specific profiles of Post-Soviet States regarding the current population density and size with proportion of population aged ≥65 years old, total expenditure on health per capita and percent of GDP, number of air passengers as well as world banking rank based on country income are presented in **Table 1**. Russia, Belarus, Kazakhstan, Azerbaijan, and Ukraine have total expenditures on health per capita more than $250 and ranked as upper middle-income countries along with Armenia and Georgia. Countries including Belarus, Georgia, Russia and Ukraine have more than 15% of their populations aged ≥65 years old.

**Table 1.** Country profiles and parameters are provided in the fitted model for each country.

| Country | Population<br>(2019),<br>mln | Population<br>density<br>person/km<sup>2</sup><br>(2019) | % of<br>population<br>65 years<br>or over<br>(2019) | Current<br>health<br>expenditure<br>per capita<br>(current<br>US\$, 2017) | Total<br>expenditure<br>on health as<br>% of GDP<br>(2017) | Air<br>passengers<br>(2018),<br>mln | Air<br>passengers<br>per capita<br>(2018) | World<br>bank<br>rank |
| --- | --- | --- | --- | --- | --- | --- | --- | --- |
| Armenia | 2.9 | 104 | 11 | 407.6 | 10.4 | 2.83 | 0.98 | UMIC |
| Azerbaijan | 10.0 | 122 | 6 | 275.8 | 6.6 | 2.28 | 0.23 | UMIC |
| Belarus | 9.4 | 47 | 15 | 342.5 | 5.9 | 2.76 | 0.29 | UMIC |
| Georgia | 4.0 | 58 | 15 | 293.0 | 7.6 | 0.52 | 0.13 | UMIC |
| Russia | 145.9 | 9 | 15 | 585.9 | 5.3 | 99.33 | 0.68 | UMIC |
| Kazakhstan | 18.5 | 7 | 8 | 279.6 | 3.1 | 7.14 | 0.39 | UMIC |
| Kyrgyzstan | 6.4 | 33 | 5 | 78.8 | 6.2 | 0.71 | 0.11 | LMIC |
| Moldova | 4.0 | 123 | 12 | 191.2 | 7.0 | 1.14 | 0.29 | LMIC |
| Ukraine | 44.0 | 76 | 17 | 177.4 | 7.0 | 7.85 | 0.18 | LMIC |
| Uzbekistan | 33.0 | 78 | 5 | 98.8 | 6.4 | 3.06 | 0.09 | LMIC |

### COVID-19 outbreak timeline and taken state preventive measures

Outbreak dates with implementation of preventive measures by country are presented in **Table 2**. Russia reported the first confirmed case of COVID-19 infection (January 31, 2020), which was earlier than other Post-Soviet States. At the end of February 2020, Azerbaijan, Belarus, Georgia, Armenia and Ukraine declared the first positive cases in their territories. Kazakhstan, Uzbekistan and Kyrgyzstan reported their first cases approximately two weeks later. Prior to introducing a state emergency and imposing lockdowns, countries implemented similar measures to prevent spread of COVID-19 by limiting air travels to countries affected by the pandemic, closure of educational institutions and banning all mass gathering events (**Table 2**). Kazakhstan, Kyrgyzstan, Uzbekistan and Moldova were quick to introduce state emergency and closed their borders, whereas the Russian Federation waited significantly longer (two months after the first confirmed case). On the other hand, Belarus, as for April 18, 2020, has not yet imposed any preventative measures such as a state emergency, or national lockdown to prevent further spread of COVID-19.

**Table 2.** Important outbreak dates and implement preventive measures by country.

| **Country** | **Date of first confirmed case in country (2020)** | **Days after first confirmed case in China** | **Actions taken prior declaration of state emergency** | **State emergency introduced days after first confirmed case in country** |
| --- | --- | --- | --- | --- |
| Armenia | 1 Mar | 54 | Prohibited entrance of citizens from countries affected by the pandemic | 15 |
| Azerbaijan | 28 Feb | 52 | Closure of borders, of educational institutions, suspension of all mass gathering events | 32 |
| Belarus | 28 Feb | 52 | Self-quarantine for 14 days after arrival, the spring vacation was extended in schools | not yet |
| Georgia | 26 Feb | 50 | Screening people travelling from abroad and limiting air connections, prohibited entrance of foreign nationals and closure of educational institutions. | 24 |
| Russia | 31 Jan | 24 | Restricting the border with countries affected by the pandemic and limiting air connections | 59 |
| Kazakhstan | 13 Mar | 66 | Screening people travelling from abroad and limiting air connections, | 3 |
| Kyrgyzstan | 18 Mar | 71 | All borders to foreigners were closed on Mar 18 | 4 |
| Moldova | 7 Mar | 60 | Closure of all kindergartens, schools, colleges and universities | 10 |
| Ukraine | 3 Mar | 56 | Limiting air travelling to epidemic places, closure of educational institutions, banning gatherings of over 200 people and cancelling flights to countries with unfavorable epidemic situations | 22 |
| Uzbekistan | 15 Mar | 68 | Limiting air connections with countries most affected by the pandemic, closure of entertainment venues and banning large weddings and other mass gatherings | 8 |

Source: official websites of Ministries of Health

### Mathematical modeling of COVID-19 outbreak

The mathematical modeling of COVID-19 including current number of confirmed, recovered and deceased cases as well as their forecasting for 10 days presented in **Figure 2**. The model predicted that the number of active new cases would be decreasing during the upcoming week in Armenia only. On the other hand, it is estimated that Kazakhstan, Kyrgyzstan, Azerbaijan and Moldova are likely to reach the epidemic peak in the upcoming weeks. Uzbekistan, Russia, Belarus, Georgia and Ukraine have the steepest increase in the number of cases and are expected to continuously increase in the next 10 days.

Based on the forecasting modelling for 30 days (**Figure 3**), similar trends expected for Kyrgyzstan, Armenia, Azerbaijan, Kazakhstan and Moldova with maximum number of approximately 650, 800, 1,000, 2,100, and 2,500 active cases (except recovered and death cases) respectively, during the peak of the worst-case scenario. In the same case scenario, the rest of the countries would not reach the peak until mid-May with maximum numbers reaching approximately 700, 5,000, 53,000 144,000 and 800,000 active cases (except recovered and death cases) for Georgia, Uzbekistan, Ukraine, Belarus, and Russia, respectively.

Increasing the protection rate for Belarus, Russia, and Ukraine would likely have significantly effects on the overall curve of active cases (**Figure 3**). The countries that were predicted to expect the most significant decrease due to strengthening of quarantine measures were Belarus and Russia.

Based on the projection for 60 days of the worst-case scenario, Russia, Belarus, and Ukraine were shown to be at risk of prolongation of the epidemic peak until mid-summer and number of active cases could be tremendously large (**Figure 4**). Armenia, Azerbaijan, and Moldova are expecting to see a decreasing trend and stabilization of the epidemic, as increasing the protection rate would reduce the number of active cases and delay the epidemic peak for these countries. Kazakhstan, Kyrgyzstan, and Uzbekistan seem to be expecting decreasing trend of active cases by the end of 60 days, in the case of maintaining the current protection rate at same level. Belarus, Russia, and Ukraine could benefit from tightening quarantine measures (increasing the protection) in the coming 60 days, which would substantially decrease the number of active cases after peak, while Armenia, Azerbaijan, Georgia, Kazakhstan, Kyrgyzstan, Moldova and Uzbekistan are unlikely to benefit from any further increase in the protection rate.

Based on current modeling approach the approximate total number of COVID-19 infected patients in both best- and worst-case scenarios from the April 18, 2020 until forthcoming 30 days and 60 days, are presented in **Supplementary Table 2**.

**Figure 2.**
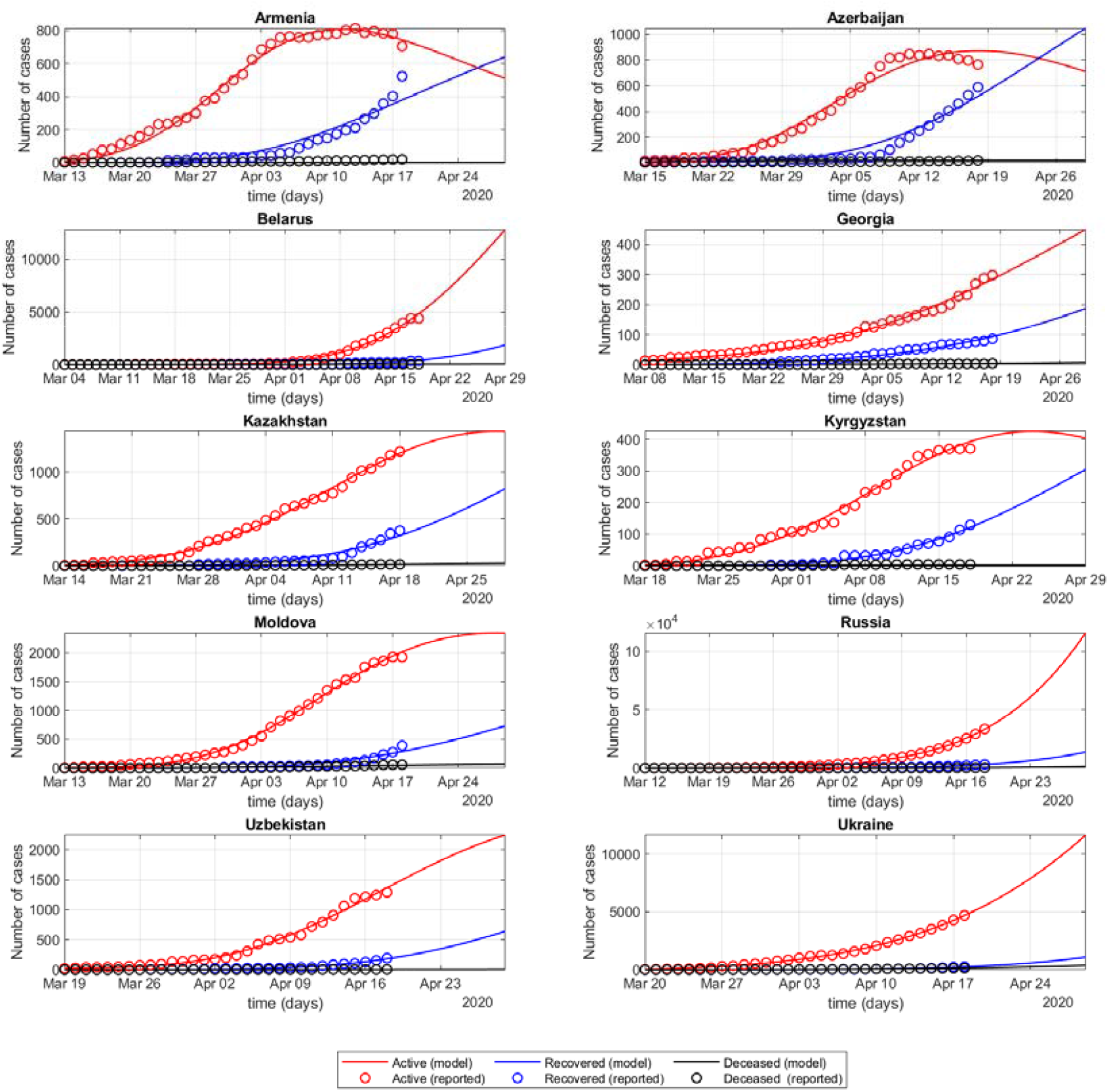
Modeling of COVID-19 outbreak prediction for 10 days. Note: Active (model) – mathematical modeling of active cases, which are all confirmed cases, excluding recovered and died; Active (reported) – laboratory confirmed active cases, which are all confirmed cases, excluding recovered and died, over time based on reported data; Recovered (model) – mathematical modeling of recovered cases; Recovered (reported) – recovered cases over time based on reported data; Deceased (model) – mathematical modeling of COVID-19 deaths; Deceased (reported) – number of deaths over time based on reported data.

**Figure 3.**
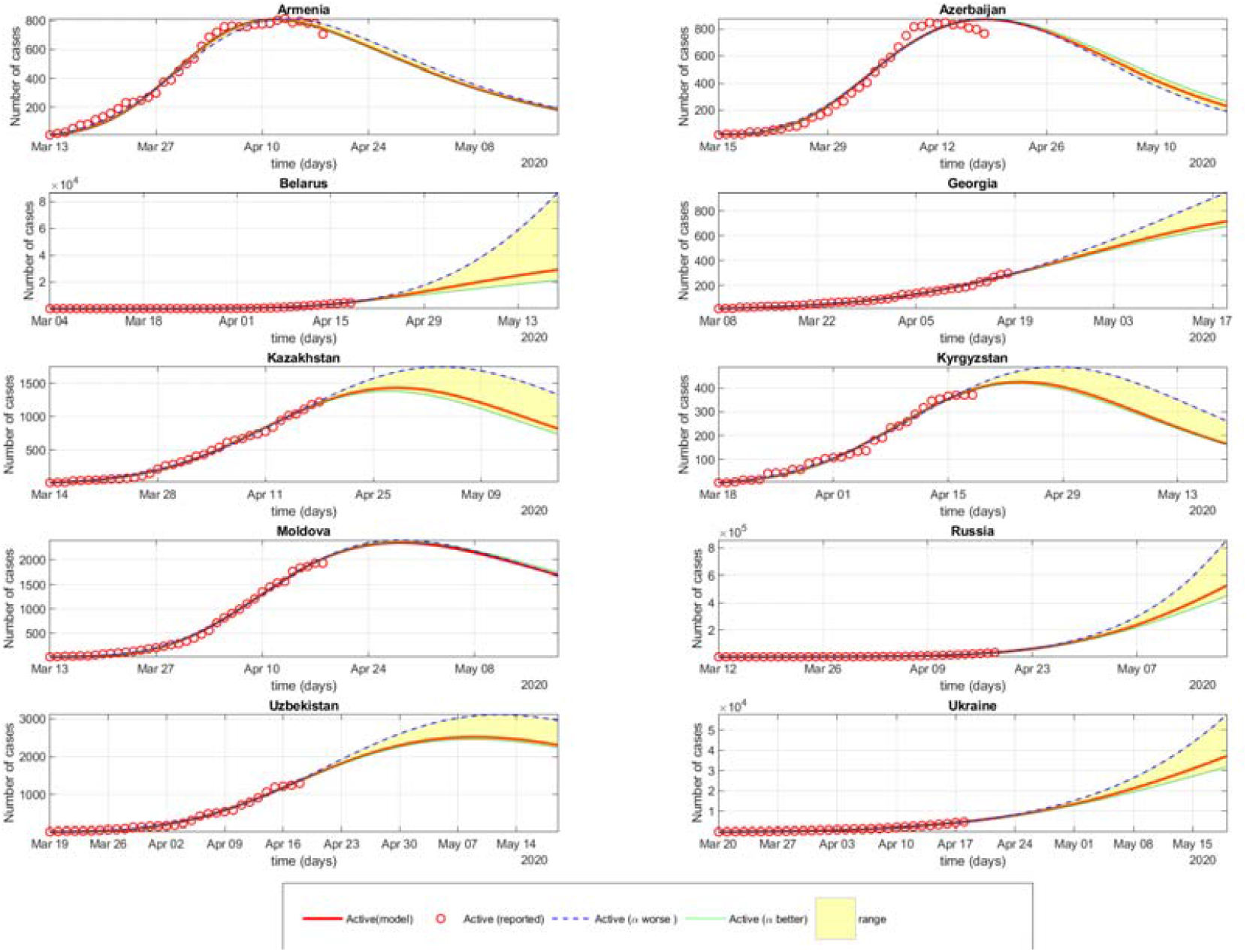
Better and worse spread scenarios of COVID-19 outbreak for 30 days.

**Figure 4.**
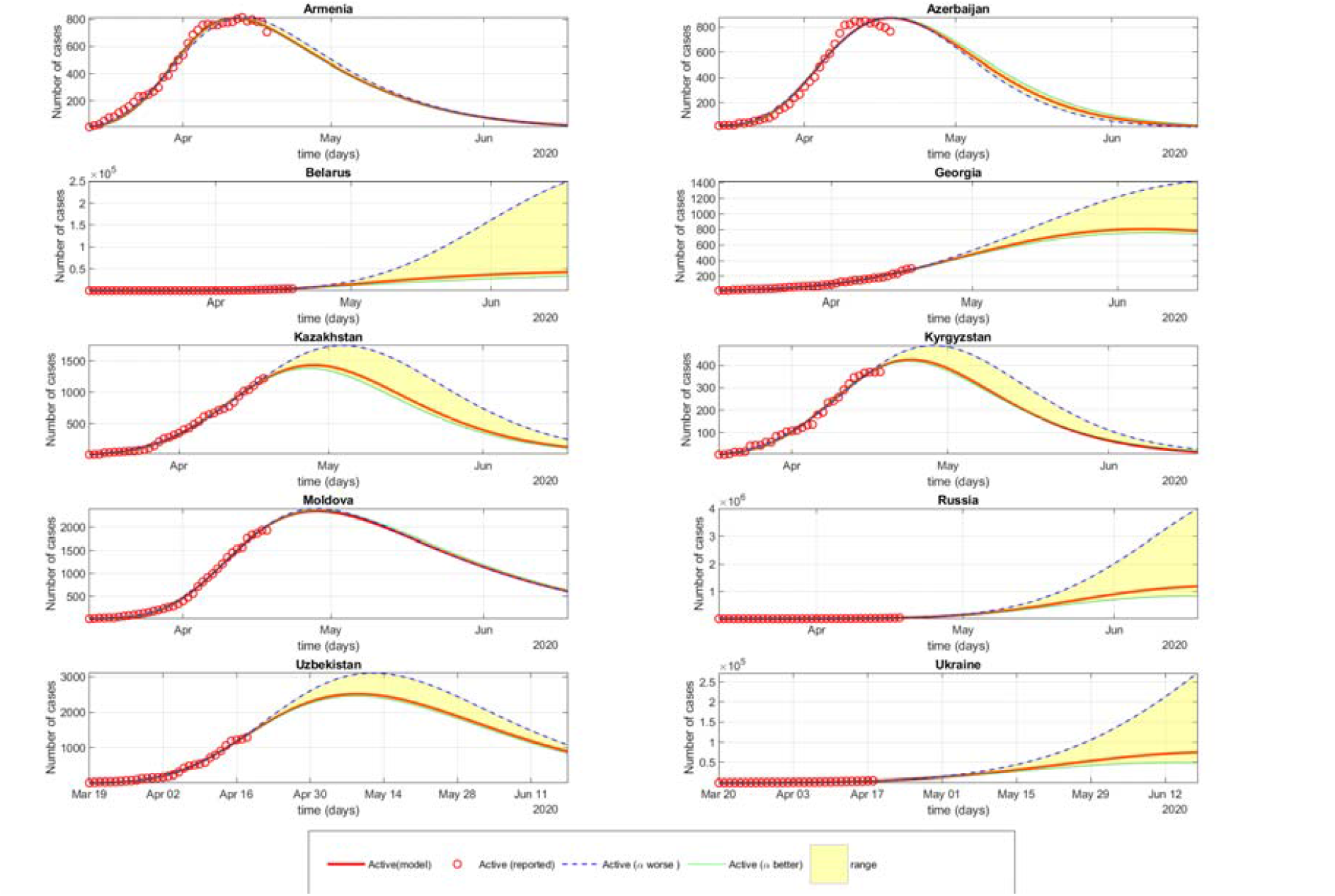
Better and worse spread scenarios of COVID-19 outbreak for 60 days. Note: Active (model) – mathematical modeling of active cases, which are all confirmed cases, excluding recovered and died; Active (reported) – laboratory confirmed active cases, which are all confirmed cases, excluding recovered and died, over time based on reported data; Active (α worse) – mathematical modeling of active cases over time for the possible worst case scenario, in which protection rate was decreased by 50%; Active (α better) – mathematical projection of active cases over time for the possible best-case scenario, protection rate was increased by 20%; range – an interval which includes all possible active cases between the worst- and best-case scenarios.

## DISCUSSION

In the current study we discuss the results of the deterministic mathematical modeling of spread of COVID-19 in the post-Soviet countries using existing data and the potential of best- and worst-case scenarios for the pandemic in each country individually. It was estimated that R_0_ value is steeply decreasing in all countries suggesting that the governments have implemented more stringent measures to prevent the spread of COVID-19. The estimated R_0_ for Belarus was the highest out of all other countries highlighting the Belarusian government’s reluctance to take stricter actions to battle COVID-19 outbreak. On the other hand, Kazakhstan, and Armenia were showing promising R_0_ values less than <1 indicating a potential reduction in transmissibility in the upcoming weeks. However, due to insufficient data in early stages of the outbreaks, estimated R_0_ values should be recalibrated for Belarus as it might not accurately predict the future dynamics of COVID-19.

Despite the relatively similar dates of the first confirmed cases across the post-Soviet countries (13), the predictions for COVID-19 outbreak dynamics in each country were significantly different. For instance, Armenia, Moldova, Georgia, Uzbekistan, Kyrgyzstan, Azerbaijan, and Kazakhstan, would likely have relatively a smaller number of cases than Russia, Ukraine, and Belarus, suggesting the success of the early governmental imposed prevention measures.

It should also be noticed that the timeline of the outbreaks is associated with proximity to the epidemic of European countries (29). The Central Asian countries (Kazakhstan, Kyrgyzstan and Uzbekistan) have faced the outbreaks later and almost at the same time and were quick to implement national preventive measures. Geographical remoteness and low air travelling from the epidemic European countries to the Central Asian countries could have delayed epidemic introduction and allowed them to learn and act more effectively than others (22, 23). On the other hand, the significantly large expected numbers of COVID-19 cases in Belarus, Ukraine and Russia could put a substantial burden on the healthcare systems in these countries. Given the comparatively larger percentage of population older than 65 years old in these countries, it could be anticipated that older individuals with COVID-19 would overwhelm ICU capacity in all hospitals in the major cities which may lead to high mortality (18). Considering low total expenditure on health as percentage of GDP and reluctance from the Belarusian government to implement similar stringent preventive measures, the state could be in a disastrous situation where all healthcare resources could be exhausted to treat the unbearably large number of severe cases (21).

Containment of epidemics in inherently weak health care systems of the Post-Soviet States could be quite a challenging task to deal with. Despite the seemingly successful containment efforts in some countries, there is a possibility of repeated epidemics after loosening quarantine measures since a large proportion of the population will be susceptible to COVID-19 or until an effective prophylaxis or post infection treatment is developed and manufactured at population scale.

Post-Soviet States have considered the economic situation and preparedness of their healthcare systems in handling the current epidemic, and consequently have taken similar and stringent actions to prevent the spread, except Belarus (6, 7). The results of current modeling show that Armenia, Azerbaijan, Georgia, Kazakhstan, Kyrgyzstan, Moldova and Uzbekistan have taken effective preventive actions against the COVID-19 spread. However, Belarus with a weak economy and unwillingness to slow down the economy could be facing extreme consequences from potential collapse of their healthcare system as well as to possibly become one of the states with the largest number of COVID-19 casualties. Ukraine and Russia were slow in placing preventive actions from the beginning of the outbreak, and without abiding stringent actions they could be facing extreme number of cases comparable to some Western European countries. The Central Asian countries will most likely fortunate in avoiding major catastrophe with the current outbreak of COVID-19, which is partly due to their early preparation for the outbreak as well as their geographical remoteness and access for international travel which played a role in relatively delayed first cases in these countries.

## Limitations

There are several limitations in this study. First, there could be some inaccuracies between reported data and what are the actual numbers are due to limited testing, reporting, and the potential unreported number of asymptomatic virus infected individuals. This, however, is a common limitation given the novelty of the virus and limited access to testing in the majority of countries. The second limitation is that some countries have implemented certain policies that limit the reporting of incidence of COVID-19, and therefore, we cannot provide predictions for Turkmenistan and Tajikistan. The third limitation is that the current burden of COVID-19 is comparatively in the early stages of development and somewhat insignificant in some post-Soviet countries, which makes our projections less robust. The fourth limitation is that we do not take into account the spread within a specific country (regional, geographical), since we considered countries as entities, and individuals are not considered. For instance, in Kazakhstan, first cases and the majority of cases were reported in densely populated major cities (Almaty and Nur-Sultan), while the outbreak reached other less populated cities a week later. The further limitation is that the modeling is not stratified by age; thus, we could not estimate the number of cases of whom would likely to require ICU treatment. Another limitation is that we do not consider how each of preventive measures (e.g. wearing masks, physical distancing, hand washing) plays role in the protection rate and dynamics of the spread within healthcare settings. In addition, mathematical models have a common limitation; they might inaccurately predict future realistic data due to insufficient data for some countries.

Nonetheless, given the limitations, we attempted to provide insights into the future possible scenarios for policy makers and decision makers about the importance of timely actions and the possible consequences of relaxing the implemented measures.

## Conclusions

Government responses were shown to be the major single factor in determining the rate of development of the COVID-19 epidemic in Post-Soviet States. Our model shows that the implementation of strict preventative measures can substantially reduce the spread of COVID-19 and that the premature loosening of these measures, in the worst-case scenario, could lead to a dramatically increase in the number of active cases and a possible prolongation of the epidemic. Based on the current confirmed cases, our model suggested that Russia, Belarus, and Ukraine would have higher cases than other Post-Soviet countries if preventive actions were to be relaxed. The estimated possible scenarios based on the proposed model can potentially be used by healthcare professionals from each studied Post-Soviet States as well as others to improve plans to contain the current and future epidemic.

## Data Availability

No Data

## Acknowledgement

YA would like to acknowledge support of the FDCRG (Nazarbayev University), No 110119FD4502, the research grant, No AP08052762, from the Ministry of Education and Science of the Republic of Kazakhstan and the SPG (Nazarbayev University). AK was supported in part by Nazarbayev University FDCRG N 090118FD5353. AG acknowledge the funding from the Nazarbayev University Collaborative Research Program (CRP) for 2020 -2022 (Funder Project Reference: 091019CRP2105) and Faculty Development Research Grant Program FDCRGP 2020-2022 (Funder Project Reference: 240919FD3913).

## SUPPLEMENTARY MATERIAL

### Appendix 1. SPEIQRD framework

In the SPEIQRD framework where a country population of size, N, is divided into seven subclasses such as S, Susceptible population: these are the individuals who have never been exposed to the COVID-19 virus; P, Insusceptible population: this is a subpopulation of S which contains individuals who become insusceptible due to a protection; E, Exposed population: asymptomatic individuals who have been in contact with COVID-19 agent but do not transmit the disease (in a latent period); I, Infected population: infected individuals can transmit the disease; Q, Isolated or quarantined population: this is a subpopulation of I which was isolated or quarantined due to being tested COVID-19 positive; R, Recovered population: a subpopulation of Q individuals who have ceased being infectious; D, Removed (dead) population: a subpopulation of Q individuals dead due to COVID-19 infection

The total population satisfies the equation N = S(t) + P(t) + E(t) + I(t) + Q(t) + R(t)+D(t). We refer to a schematic illustration of the model in the figure which takes the following form Model 1:

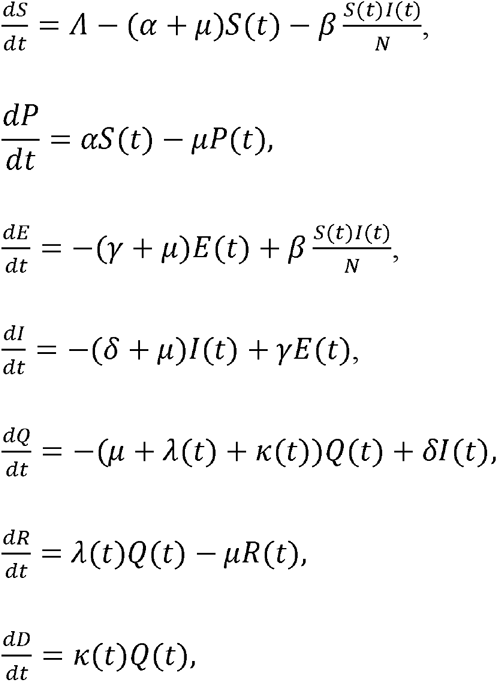

Where 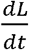 models the rate of change in the subpopulation L=(30 I, Q, R, D) at day t, *⋀* is the rate at which individuals are born into the population *α* is the protection rate, µ is the natural death rate, β is the per capita infection rate of an average susceptible provided that everyone else is infected, γ and δ are the rates at which individuals leave the exposed and infectious classes, respectively. Moreover, *λ*(*t*) and *k*(t) are the treatment and disease-related (COVID-19) death rate, respectively. Thus, 1/µ is the average life expectancy, 1/N gives the probability that a given contact is with an infectious individual. Further, 1/γ and 1/δ give the average length of the latent and isolation (quarantine) periods, respectively. For the treatment and COVID-19 death rates we have used the following time-dependent functions: *κ*(t)= *κ*_1_*exp(-*κ*_2_*t) and *λ* (t) = *λ*1 * (1 - exp(-*λ*2 * t)).

The parameters are listed below.

*⋀* the rate at which individuals are born into the population

*α* the protection rate

*β* the per capita transmission rate of the disease

*λ*(*t*) the treatment rate

*κ*(*t*) the disease-related (COVID-19) death rate

1/*µ* the average life expectancy

1/*γ* the average length of the latent period

1/*δ* the average length of the isolation (quarantine) period

Once we have a model the next question is to determine in terms of the given parameters so-called basic reproduction number, **R**_**0**_, which serves as the threshold value. In epidemiology, **R**_**0**_ is defined as the average number of new infectious cases produced by an infectious member of the host population. It is known that if **R**_**0**_>1 then there will be an outbreak, otherwise, if **R**_**0**_<1, the disease will die out. Thus, it is of utmost importance to find **R**_**0**_. Note that the average length of infection is 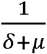 and recall that *β*is the per capita infection rate. Further, the probability of an individual in exposed population becoming infectious before dying is 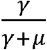. Thus, in the case of *α*=0, the basic reproduction number can be computed as:

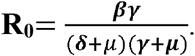

In the Covid-19 case, we take into account that the susceptible populations are decreasing not only due to infection and death but also becoming insusceptible due to protection measures such as lock-downs of cities and/or contact tracing of individuals. Therefore, *α* ≠ 0 in our model. In this case, following the study from [1] we compute the basic reproduction number as

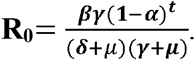

---

**Supplement Table 1.** Parameters are provided in the fitted model for each country as April 18, 2020.

| Country | $\alpha$ | $\beta$ | $\delta$ | $\gamma$ | $\lambda_1$ | $\lambda_2$ | $k_1$ | $k_2$ | $R_0$ |
| --- | --- | --- | --- | --- | --- | --- | --- | --- | --- |
| Armenia | 4.28E-02 | 1.1503 | 0.555 | 0.8514 | 0.4694 | 0.002 | 0.0024 | 1.8897 | 0.41 |
| Azerbaijan | 1.52E-01 | 1.4704 | 0.0766 | 0.2405 | 0.84 | 0.0019 | 0.2501 | 0.4101 | 0.061 |
| Belarus | 5.07E-02 | 1.1438 | 0.0015 | 0.3645 | 0.0289 | 0.0149 | 0.6999 | 0.1821 | 68.4017 |
| Georgia | 3.17E-02 | 1.0829 | 0.1722 | 0.0464 | 0.0233 | 0.1832 | 0.0009 | 0 | 1.6217 |
| Kazakhstan | 1.60E-02 | 1.127 | 0.6788 | 0.7002 | 0.4386 | 0.0019 | 0.0015 | 0.011 | 0.9305 |
| Kyrgyzstan | 2.03E-02 | 1.2222 | 0.7359 | 0.7339 | 0.4083 | 0.0028 | 0.2824 | 1.8208 | 0.8612 |
| Moldova | 7.67E-02 | 1.2833 | 0.163 | 0.3382 | 0.2922 | 0.0015 | 0.0131 | 0.0687 | 0.411 |
| Russia | 1.16E-02 | 1.0165 | 0.4133 | 0.403 | 0.0142 | 0.1174 | 0.0014 | 0.0001 | 1.5774 |
| Uzbekistan | 8.92E-02 | 1.0823 | 0.0731 | 0.2132 | 0.1213 | 0.0058 | 0.0026 | 0.0933 | 0.817 |
| Ukraine | 3.55E-03 | 1.1329 | 0.8715 | 1 | 0.0623 | 0.0049 | 0.005 | 0.0177 | 1.1683 |

*Abbreviations: $\alpha$ – alpha; $\beta$ – beta; $\gamma$ – gamma; $\delta$ – delta; $\kappa$ – kappa; $R_0$ – basic reproduction number.*

Based on the mathematical modelling (**Supplement Figure 1**) it was estimated that among post-Soviet countries, Azerbaijan, Armenia, Moldova, Uzbekistan, Kyrgyzstan and Kazakhstan have managed to lower the reproduction number (R_0_) <1 (0.061, 0.41, 0.411, 0.817, 0.8612 and 0.9305, respectively). In the rest of the states, the reproduction numbers were quite close to 1, not exceeding 1.63 (Russian Federation and Uzbekistan). Belarus had the largest and extreme reproduction number (R_0_=68.4).

**Supplementary Table 2.**
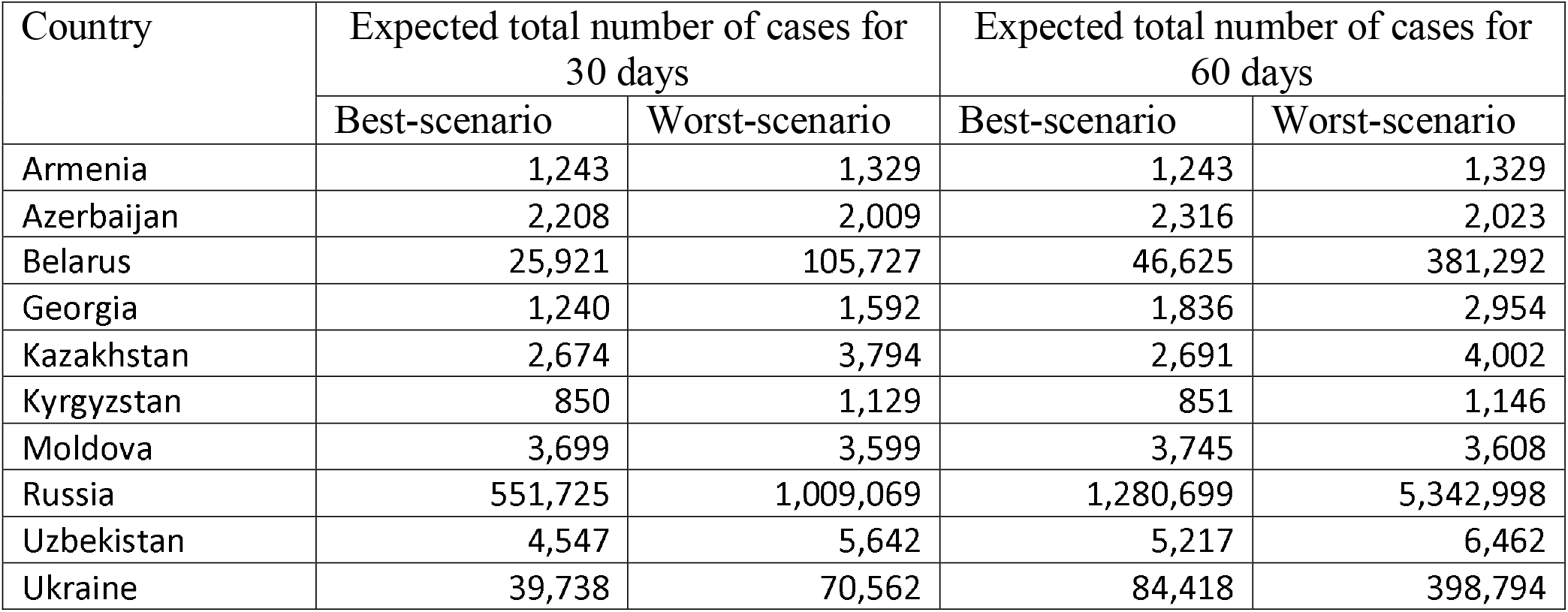
Expected total number of COVID-19 confirmed cases in both best- and worst-case scenarios from the April 18, 2020 until forthcoming 30 days and 60 days.

| Country | Expected total number of cases for<br>30 days |  | Expected total number of cases for<br>60 days |  |
| --- | --- | --- | --- | --- |
|  | Best-scenario | Worst-scenario | Best-scenario | Worst-scenario |
| Armenia | 1,243 | 1,329 | 1,243 | 1,329 |
| Azerbaijan | 2,208 | 2,009 | 2,316 | 2,023 |
| Belarus | 25,921 | 105,727 | 46,625 | 381,292 |
| Georgia | 1,240 | 1,592 | 1,836 | 2,954 |
| Kazakhstan | 2,674 | 3,794 | 2,691 | 4,002 |
| Kyrgyzstan | 850 | 1,129 | 851 | 1,146 |
| Moldova | 3,699 | 3,599 | 3,745 | 3,608 |
| Russia | 551,725 | 1,009,069 | 1,280,699 | 5,342,998 |
| Uzbekistan | 4,547 | 5,642 | 5,217 | 6,462 |
| Ukraine | 39,738 | 70,562 | 84,418 | 398,794 |

**Supplement Figure 1.**
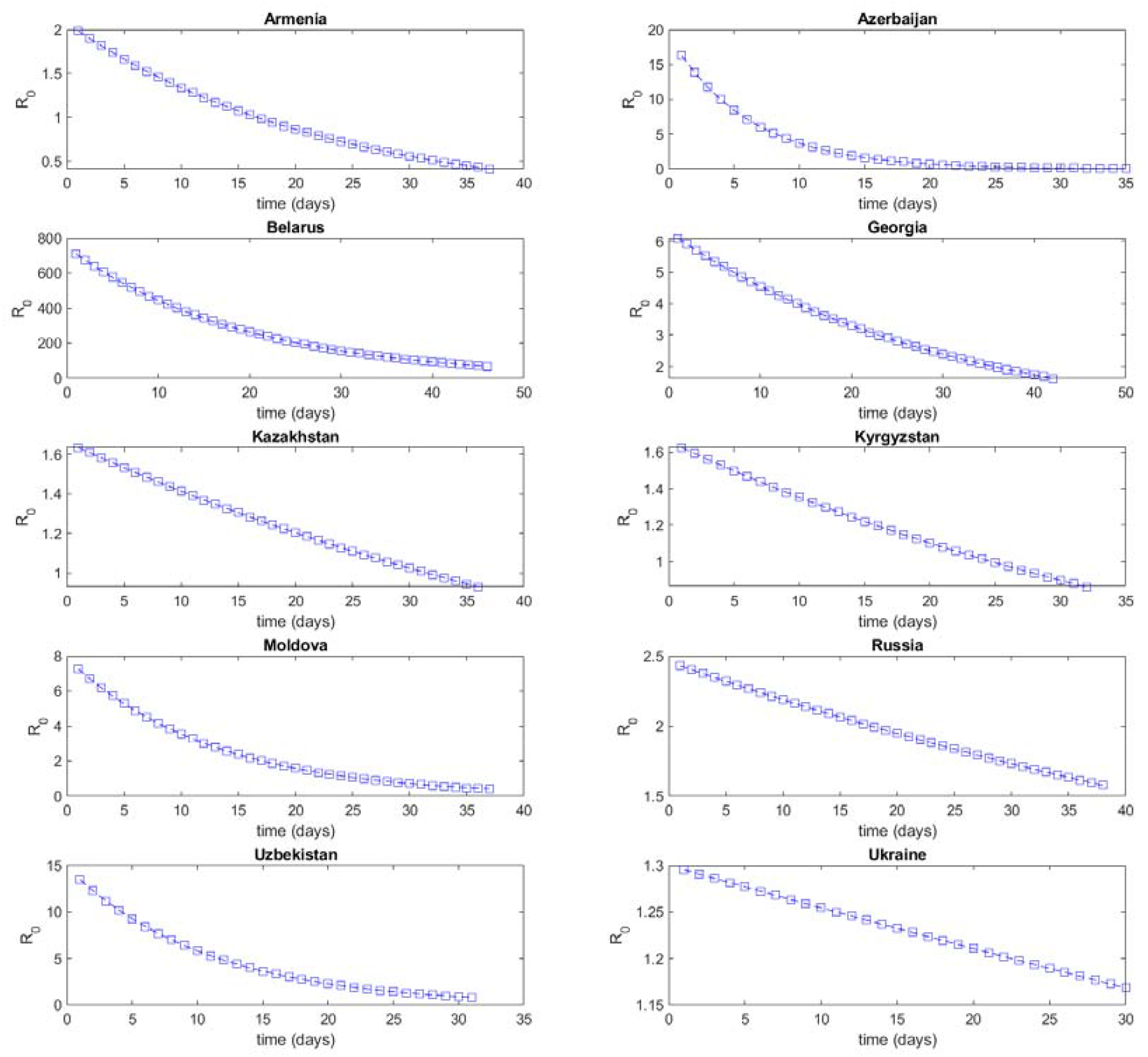
R_0_ in modeling of COVID-19 outbreak for post-soviet countries. *Abbreviations: R*_*0*_ *– reproduction number*.

